# An integrated active case search for skin-NTDs in yaws endemic health districts in Cameroon, Côte d’Ivoire and Ghana

**DOI:** 10.1101/2023.11.16.23298508

**Authors:** Serges Tchatchouang, Laud A. Basing, Hugues Kouadio-Aboh, Becca L. Handley, Camila G-Beiras, Ivy Amanor, Philippe Ndzomo, Mohammed Bakheit, Lisa Becherer, Sascha Knauf, Claudia Müller, Earnest Njih-Tabah, Theophilus Njmanshi, Tania Crucitti, Nadine Borst, Simone Lüert, Sieghard Frischmann, Helena Gmoser, Emelie Landmann, Aboubacar Sylla, Mireille S. Kouamé-Sina, Daniel Arhinful, Patrick Awondo, Gely Menguena, Emma-Michèle Harding-Esch, Adingra Tano, Mamadou Kaloga, Paul Koffi-Aboa, Nana Konama-Kotey, Oriol Mitjà, Sara Eyangoh, Kennedy Kwasi-Addo, Solange Ngazoa-Kakou, Michael Marks

## Abstract

**Background:** Integrated approaches to mapping skin Neglected Tropical Diseases (NTDs) may be cost-effective way to guide decisions on resource mobilization. Pilot studies have been carried out, but large-scale data covering multiple countries endemic for skin-NTDs are lacking. Within the LAMP4YAWS project, we collected integrated data on the burden of multiple skin-NTDs.

**Methods:** From March 2021 to March 2023, integrated case searches for yaws alongside other skin conditions were performed in endemic health districts of yaws in Cameroon, Côte d’Ivoire, and Ghana. Initial screening involved a brief clinical examination of participants to determine if any skin conditions were suspected. Cases of skin-NTDs were then referred to a health facility for appropriate management.

**Results:** Overall 61,080 individuals screened, 11,387 (18.6%) had skin lesions. The majority of individuals (>90%) examined were children aged 15 years old and under. The proportion of serologically confirmed yaws cases was 8.6% (18/210) in Cameroon, 6.8% (84/1232) in Côte d’Ivoire, and 26.8% (440/1643) in Ghana. Other skin conditions based on clinical examination included: scabies, Buruli ulcer, leprosy, lymphatic filariasis (lymphoedema and hydrocele), tungiasis, and fungal infections. The most common conditions were scabies and superficial fungal infections (scabies versus fungal infections) in Cameroon with 5.1% (214/4204) versus 88.7% (3730/4204), Côte d’Ivoire with 25.2% (1285/5095) versus 50.4% (2567/5095) and Ghana 20% (419/2090) versus 1.3% (28/2090). Other skin-NTDs were less common across all three countries.

**Conclusion:** This study confirms that integrated screening allows simultaneous detection of multiple skin-NTDs, maximising use of scarce resources.

**Plain English Summary:** Many Neglected Tropical Diseases (NTDs) predominantly affect the skin and are referred to as skin-NTDs. The World Health Organization (WHO) has developed a number of strategies for the control, eradication and elimination of skin-NTDs and recognizes the importance of integrated approaches to mapping skin-NTDs. We conducted a study adopting integrated screening for multiple skin-NTDs and other skin conditions in Cameroon, Côte d’Ivoire, and Ghana. This ran alongside a study focused on diagnostic tests for one specific skin-NTD – yaws.

The results showed that integrated screening is a feasible and cost-effective way to detect multiple skin-NTDs in a single intervention. Of more than 60,000 individuals screened almost one in five had a skin lesion. Cases of yaws confirmed by blood tests were detected more frequently in Ghana compared to Côte d’Ivoire and Cameroon. The most common skin conditions were scabies and superficial fungal infections. Other skin-NTDs such as Buruli ulcer, leprosy, lymphatic filariasis and tungiasis were less common. Integrated screening allowed detection of skin conditions and co-endemicity of skin-NTDs and the data can guide decisions on resource mobilization to manage skin-NTDs.

## INTRODUCTION

Skin diseases affect over 900 million people worldwide annually and are the third most frequent reason for seeking care (1–3). A subset of these conditions are also classified by the World Health Organization (WHO) as Neglected Tropical Diseases (NTDs) (4). Of the twenty NTDs recognised by WHO, more than half can present as skin diseases and are referred to as skin-NTDs. If not diagnosed and treated promptly, these conditions can result in disfigurement, disability, stigmatisation, impacts on mental health, and socioeconomic difficulties (4–6).

In Cameroon, Côte d’Ivoire, and Ghana, several skin-NTDs are known to be endemic including yaws (7–10), Buruli ulcer (11–13), leishmaniasis (14–16), leprosy (10, 17, 18), lymphatic filariasis (19–21), mycetoma (22–24), onchocerciasis (25–27), and scabies (10, 28–30). Alongside these conditions, a number of more common skin conditions including superficial fungal and bacterial infections cause a considerable burden of disease (28, 31–33).

Most data on skin-NTDs rely on passive surveillance activities, which are likely to underestimate the true burden of disease as they are dependent on patients who present to a health centre, receive a diagnosis and are reported to the respective disease control programme. The highest burden of skin-NTDs is believed to be in communities living in difficult to reach areas with limited access to healthcare (34, 35). Consequently, routine data are likely to underreport the true burden and epidemiology of these conditions.

To achieve the WHO goals for skin-NTDs, active case detection will be required. Therefore, WHO has recently promoted integrated activities of skin-NTDs in order to accelerate progress in their control, elimination and eradication (4, 36). Because NTDs often overlap in geographic areas, integrated active case searching may offer a way to overcome some of the challenges associated with stand-alone programmes and reduce the risk of late detection of skin-NTDs. Integration can also result in a reduction of resources, thereby increasing cost-effectiveness and allow for an expanded coverage of interventions (2, 36–38).

The LAMP4YAWS project in Cameroon, Côte d’Ivoire, and Ghana aimed to implement and evaluate a loop-mediated isothermal amplification test for yaws. As part of the project, field activities included active case searching of yaws using an integrated approach (39). Here we report data on skin conditions from active case searches in yaws-endemic health districts from all three countries.

## METHODS

### Study design

From March 2021 to March 2023, we undertook integrated case searches in rural communities within suspected yaws-endemic health districts in Cameroon, Côte d’Ivoire, and Ghana. Integrated approach for skin-NTDs offers possibility to address several diseases simultaneously in the same communities in order to maximize the utilization of limited resources (40). This approach was implemented by assessing burden of yaws in health districts from notifications of NTD national programs, training of health workers and village volunteers, conducting active case detection of skin conditions, clinical diagnosis and rapid tests for yaws, treating yaws cases and referring other skin conditions to healthcare centres (2). In each community visited, initial screening involved an examination to determine if participants had skin lesions as described in the study protocol (39).

### Study sites

The study was conducted in rural areas in 12 health districts in Cameroon, six in Côte d’Ivoire, and seven in Ghana (39). The characteristics of the study sites are summarised in Table 1. Study sites were selected based on clinical notifications of yaws by the national programmes on NTDs.

**Table 1:** Characteristic of study sites in Cameroon, Côte d’Ivoire and Ghana.

| Country | Region | Health district | Sub-divisions | Population | Active case searching strategy |
| --- | --- | --- | --- | --- | --- |
| Cameroon | East | Doumé | Doumé, Doumaitang, Dimako | 53,286 | Community, House-to-house |
|  |  | Bétaré-Oya | Betaré-Oya, Ngoura | 141,405 | Community, health centres |
|  |  | Batouri | Batouri, Ndemnam | 121,199 | Community, health centres |
|  |  | Messamena | Messamena, Somalomo | 48,000 | Community, health centres |
|  |  | Abong-Mbang | Abong-Mbang, Atok, Angossas, Dja (Mindourou) | 85,741 | Community, health centres |
|  |  | Lomié | Lomié, Messok, Ngoyla | 47,695 | Community, health centres |
|  |  | Yokadouma | Gari-Gombo, Yokadouma | 126,353 | Community, health centres |
|  |  | Mbang | Mbang | 34,148 | Community, House-to-house |
|  | South | Djoum | Djoum, Oveng, Mintom | 43,142 | MDA |
|  |  | Lolodorf | Bipindi, Lolodorf, | 43,446 | Community |
|  |  |  | Mvengue,<br>Akom 2 |  |  |
|  |  | Sangmelima | Sangmelima<br>, Meyomessi | 110,385 | School, community |
|  | Adamawa | Bankim | Bankim | 121,756 | School, community,<br>church |
| Côte<br>d'Ivoire | Loh<br>Djiboua | Divo | Goh-Djiboua | 458,318 | School, Community |
|  | Agneby<br>Tiassa | Tiassalé | Lagunes | 270,412 |  |
|  | Bélier | Toumodi | Lacs | 189,794 |  |
|  |  | Autonomou<br>s district of<br>Yamoussou<br>kro | N/A | 446,158 |  |
|  | Haut<br>sassandra | Daloa | Sassandra-Marahou | 745,628 |  |
|  |  | Vavoua | Sassandra-Marahou | 451,492 |  |
| Ghana | Central | Asikuma<br>Odoben<br>Brakwa | N/A | 129,580 | School, Community |
|  |  | Ajumako<br>Enyan<br>Essiam | N/A | 138,046 |  |
|  | Eastern | Upper West<br>Akim | N/A | 93,391 |  |
|  |  | Ayensuanor | N/A | 94,594 |  |
|  |  | Akuapim<br>North | N/A | 105,315 |  |
|  |  | Asene<br>Manso<br>Akroso | N/A | 77,498 |  |
|  | Western | Mpohor | N/A | 52,473 |  |

MDA: Mass drug administration; N/A: not applicable

### Recruitment of participants

Case searches were conducted in collaboration with national NTD programmes. District health staff were first trained to recognise skin-NTDs as well as other skin conditions. Training materials included the WHO manual on recognition of skin-NTDs (41), as well as other national NTD training materials.

Individuals presenting with yaws-like lesions underwent SD Bioline (Abbott, USA) test to detect anti-treponemal antibodies. Those reactive underwent a Chembio Dual Path Platform (DPP) test (Chembio Diagnostics, New York, USA) for simultaneous detection of both treponemal and non-treponemal antibodies, interpreted as serologically confirmed yaws case. Samples were then taken as part of the main study from participants as described elsewhere (39). Other skin diseases were recorded and referred to the health district for management following the national guidelines.

Several approaches were used for case finding, including community-based case detection, detection in healthcare centres, primary school-based screening programs, screening alongside mass drug administration (MDA) campaigns, church and house-to-house case searches (Table 1). The strategy was chosen based on other activities taking place in the selected district. The research team visited different study sites according to a pre-established schedule that was communicated to health workers, village chiefs, and school directors before the appointed day.

### Case definition of other skin conditions

In order to be considered a case of leprosy (Hansen’s Disease), an individual must have one or more of skin lesions that were hypopigmented or red and had definite loss of sensation.

Buruli ulcer case was based on clinical presentation consistent with a painless, non-healing ulcer with undermined edges and a necrotic base, typically located on the limbs; a painless nodule or plaque, typically located on the limbs; and oedema of a limb or other body part, with or without overlying ulceration.

Scabies were defined as people with rash consisting of papules, vesicles, and burrows, typically located on the hands, feet, and folds of the skin.

An individual suffering from lymphatic filariasis presented lymphedema characterised by the enlargement of body parts in the limbs and breasts; or hydrocele marked by swelling in the scrotal sack.

Case of tungiasis was defined as participant with a painful, pruritic nodule with a central black dot, mainly located on the feet, toes, or fingers.

Superficial fungal infections were defined as affection on the body commonly with a red, ring-shaped rash with a raised border for *Tinea corporis* (ringworm); scaly patches on the scalp sometime with hair loss for *Tinea capitis* (scalp ringworm); light or dark patches on the skin, most commonly on the chest and back for *Tinea versicolor* (pityriasis versicolor).

### Data analyses

Study data were recorded using standardized data collection forms. We used basic descriptive statistics to summarise the characteristics of the individuals who were screened, the number who were diagnosed with each skin-NTD, and other skin conditions. Results are shown stratified by country. Analyses were performed in Microsoft Excel 2013.

### Ethics approval

This study was approved in Cameroon by the National Ethics Committee on Research for Human Health (CNERSH) under decision N°2020/12/1327/CE/CNERSH/SP and administrative authorization of the Ministry of Public Health N°D30-308/L/MINSANTE/SG/DROS), in Côte d’Ivoire by the Comité National d’Ethique des Sciences de la Vie et de la Santé (IQR) through decision IQRG0075_16/09/2020, in Ghana by the Ghana Health Service of Ethical Review Committee (GHS-ERC) and Noguchi Memorial Institute for Medical Research-Institutional Review Board (NMIMR-IRB) under decisions GHS-ERC 005/12/20 and NMIMR-IRB CPS 019/20-21), and in United Kingdom through London School of Hygiene and Tropical Medicine (LSHTM) under decision N° 21633 (19 August 2021).

All participants and/or the parents/legal guardians of minors provided written informed consent before enrolment. In addition, assent was obtained from child participants.

## RESULTS

### Demographic characteristics

Case searches were conducted in 12 districts in Cameroon, six in Cote d’Ivoire, and seven in Ghana. A total of 61,080 individuals were screened, including 20,414 in Cameroon, 16,530 in Côte d’Ivoire, and 24,136 in Ghana. The male to female ratio was approximately 1:1 in the three countries (10804/9610, 8430/8100, and 13241/10895 respectively for Cameroon, Côte d’Ivoire and Ghana), with more than 90% of participants aged up to 15 years old (56471/61080).

### Skin lesions detected

Table 2 summarizes the skin diseases detected in Cameroon, Côte d’Ivoire, and Ghana. Skin lesion accounted for 18.6% (11389 /61080) and included 20.6% (4204/20414) in Cameroon, 30.8% (5095/16530) in Côte d’Ivoire and 8.7% (2090/24136) in Ghana.

**Table 2:**
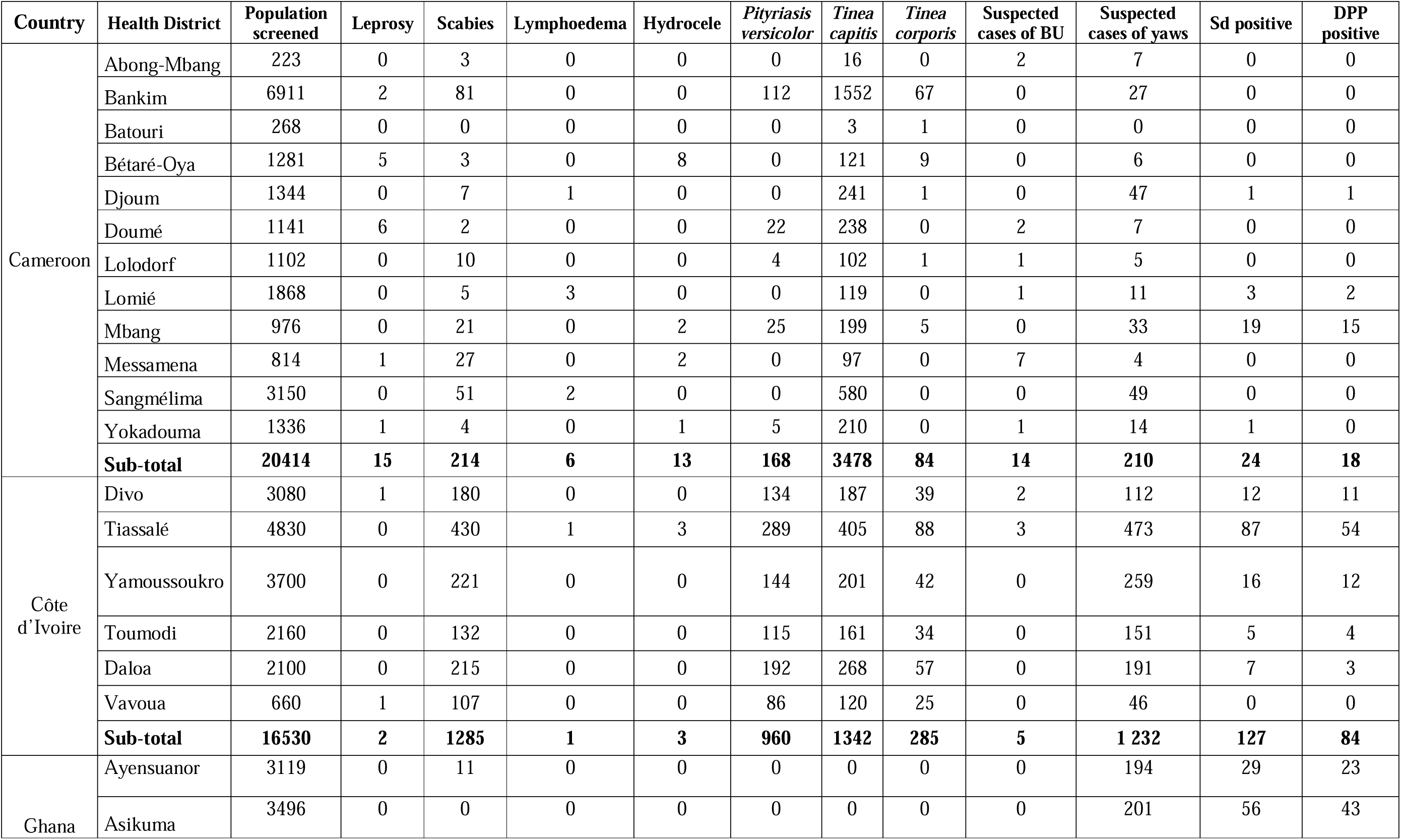
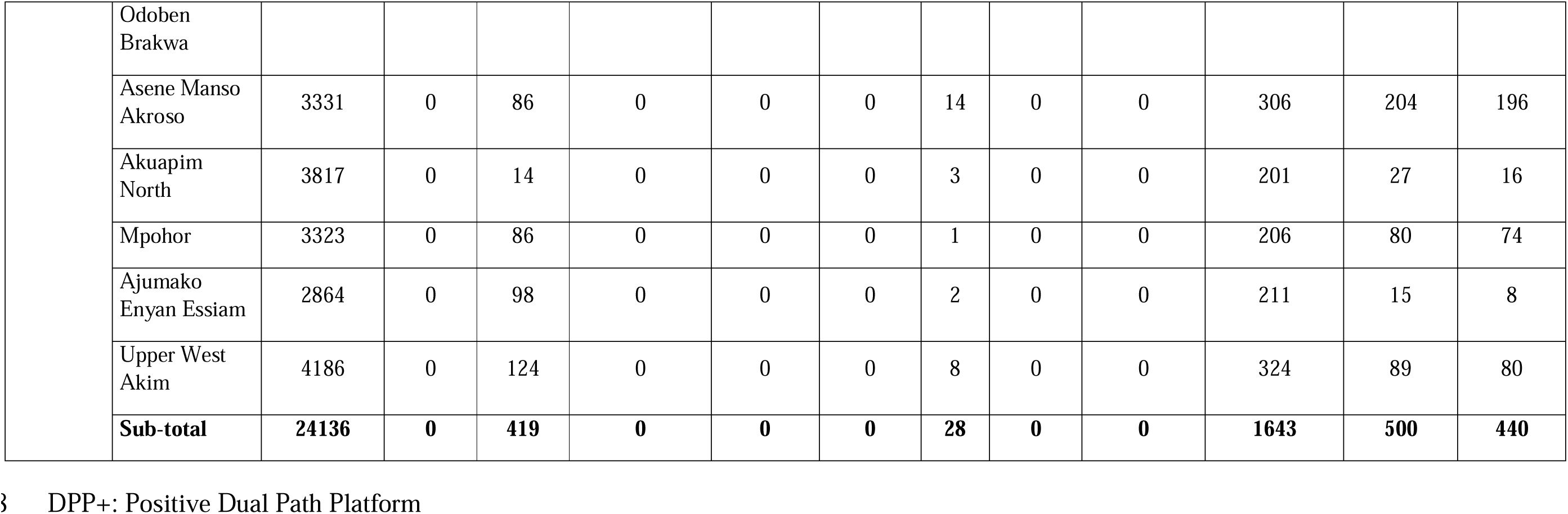
Characteristics of skin diseases found in Cameroon, Côte d’Ivoire and Ghana.

The most frequent skin conditions encountered in Cameroon were *Tinea capitis,* scabies and yaws-like lesions. *T. capitis* accounted for 82.7% (3478/4204) of the skin conditions and was detected mainly in the health districts of Bankim (44.6%, 1552/3478)) and Sangmélima (16.7%, 580/3478)). Scabies was the second most common skin condition detected in 5.1% (214/4204) cases of skin conditions seen. The majority of scabies cases were again detected in Bankim and Sangmélima with 37.9% (81/214) and 23.8% (51/214) of scabies among the detected skin lesions, respectively. Yaws-like lesions were the third most common skin condition (5%, 210/4204) and were identified in high numbers in Sangmélima (23.3%, 49/210), Djoum (22.4%, 47/210) and Mbang (15.7%, 33/210). However, only 18 individuals were serologically confirmed as yaws cases almost all among Baka population.

*Pityriasis versicolor*, scabies, and yaws-like lesions accounted for 99.8% (5084/5095) of skin conditions detected in Côte d’Ivoire. *P. versicolor* was the most common skin condition (50.4% (2567/5095)) across the six health districts visited. Scabies represented 25.2% (1285/5095) of the conditions and yaws-like lesions were reported among 24.2% of individuals presenting with a skin condition (1232/5095) and were serologically confirmed in 84 people.

In Ghana, yaws-like lesions accounted for 78.6% (1643/2090) of them, with serologically confirmed yaws among 440 individuals. Upper West Akim District and Asene Manso Akroso recorded the highest number of yaws cases. Scabies and *T. capitis* accounted for 20% (419/2090) and 1.3% (28/2090) of skin conditions, respectively.

## DISCUSSION

In accordance with the NTD road map 2021-2030 (4), skin-NTDs are targeted for eradication (yaws), elimination (leprosy, onchocerciasis, lymphatic filariasis) and control (other skin-NTDs), and an integrated approach is the backbone to achieving these targets. The main study was focused on yaws while adopting an integrated approach in the screening of individuals in the selected health districts.

This project improved case detection of skin conditions such as yaws, scabies, lymphatic filariasis, Buruli ulcer, leprosy, tungiasis and superficial fungal infections in the three countries. Yaws and scabies were the predominant skin-NTDs.

The frequency of detection of serologically confirmed yaws among suspected cases in Cameroon, Côte d’Ivoire and Ghana was 8.6%, 6.8% and 26.8% respectively. These data show that yaws is more prevalent in endemic health districts in Ghana compared to Cameroon and Côte d’Ivoire, which is consistent with routine reporting data in Ghana with frequencies above 15% among suspected cases of yaws (34, 42, 43). Ghana has the second highest population density in West Africa and this can make it more difficult to control the spread of yaws. For instance, many MDA campaigns have been performed previously but the incidence of yaws is still high in many parts of the country (42, 44, 45). The data obtained in Cameroon overlap with previous findings, with less than 10% of yaws over suspected cases (46, 47) but lower to what has been reported during an outbreak (8). The decrease of yaws cases in Cameroon is linked to the surveillance impact by identifying and treating cases, and MDA implementation in the Congo Basin in December 2020 before the current study was conducted (48). The results of Côte d’Ivoire show that yaws remains prevalent in the study sites (1.6%; 81/5095) compared to findings in the Divo health district between 2016 and 2017 that identified 8 cases of yaws out of 1,302 individuals with skin conditions (10). Côte d’Ivoire is one of the countries in Africa that has been most affected by skin-NTDs, including yaws (49, 50). The government of Côte d’Ivoire has implemented a number of programmes to address these diseases. Areas of co-endemicity of skin-NTDs have undergone several investigations with treatment which have certainly reduced the incidence of yaws (10). The results of this study suggest that additional efforts are needed to control yaws in the three countries where the disease is endemic, in particular the number of rounds of MDA which can be reviewed to significantly reduce the incidence of the disease (51). More than 80% of yaws like ulcers were not yaws (542/3085), suggesting that proper identification and management of other cutaneous ulcers, such as *H. ducreyi*, is important to improve patient outcomes.

Scabies was the most common skin-NTD detected among participants with skin conditions. The frequency was 25.2% in Côte d’Ivoire, 19.8% in Ghana and 5.1% in Cameroon. These data are high compared to previous studies in the three countries where proportions of scabies in people with skin lesions ranged almost from 3 to 9% during active case search. Difference may be due to a number of factors, including the duration of the screening, the context of the study and the methods used to detect scabies. The field activities of the current study were carried out over almost three years unlike the previous ones whose duration was over a few months. Earlier data from the three countries highlight the variability in the proportions of scabies depending on the context and methods to detect in hospital, prison, boarding school or even during active research in the community and in schools. Thus, data from several regions in Côte d’Ivoire show the proportions of scabies during active case searches among people suffering from skin diseases varied between 3.7% and 8.6% (10, 49, 52). In Ghana, recent findings from 2019 show that the proportions of scabies are between 6 to 12% among patients suffering from skin infections reported to urban and rural study health facilities (30, 53), and prevalences of 11.2% to 71% during outbreaks (54, 55). In Cameroon, surveys in urban areas in boarding schools show the prevalence of scabies of nearly 18% (56), 32% in prison (57), and a proportion of 2.8% during active case search in rural areas among respondents with skin disease (28). These data suggest that scabies is a major public health problem in all three countries.

Other skin-NTDs such as leprosy, Buruli ulcer, lymphatic filariasis and tungiasis were less common. Leprosy cases were not found in Ghana while in Cameroon and Côte d’Ivoire, leprosy cases varied between 2 and 15 throughout the study period. Available data show that some districts in Cameroon and Côte d’Ivoire are still of high endemicity (17, 31, 52). No case from Ghana could be due to the significant progress in controlling leprosy or targeted study sites. Buruli ulcer and lymphatic filariasis case numbers followed the same trend as leprosy in Cameroon and Côte d’Ivoire, and tungiasis was only recorded in Cameroon. Buruli ulcer and lymphatic filariasis are known to be endemic in Cameroon, Côte d’Ivoire and Ghana (11–13, 19–21) suggesting that the diseases may have similar risk factors, and that tungiasis may be more common in Cameroon due to specific environmental factors.

The study also found that superficial fungal infections were the most common form of skin disease in two of the three countries. In Cameroon, these conditions accounted for almost 90% with the majority due to *Tinea capitis* while *Pityriasis versicolor* constituted the bulk of these infections in Côte d’Ivoire (50.4%). The findings are concordant with field data in Cameroon, Côte d’Ivoire and Togo highlighting fungal infections as the main skin conditions (10, 28, 31, 58). In previous investigation, proportion of *Pityriasis versicolor* and *Tinea capitis* among diagnosed skin diseases in Côte d’Ivoire was approximately 45% of each (31); while in Cameroon proportion of *Pityriasis versicolor* in a rural setting was around 21% (28). In Ghana, superficial fungal infections were likely to be less common. Previous data indicated that fungal infections accounted for between 20 and 30% of patients with skin diseases (59, 60). In this study, superficial fungal infections were not automatically recorded in Ghana, thus, only *Tinea capitis* was detected in 1.3% of individuals. In previous studies in Ghana, *Tinea capitis* was one of the main fungal agents with proportions above 20% (59).

We found that the proportion of people with skin lesions was not the same from one country to another and according to the diseases. This is likely due to a variety of factors, such as ecological zones (climate, vegetation and culture), population density and standard of life. However, early diagnosis and treatment are important to prevent the spread and to reduce the risk of complications. Reducing the burden of skin conditions may require improved access to clean water and sanitation facilities, education of people about the diseases and their prevention measures, availability and affordability of treatment, and reinforcement of public health measures to control the spread of diseases such as mass drug administration programmes.

All data were recorded at the health district level and subsequently transmitted to the respective NTD programmes. Through these data, we can see the diversity that can exist in terms of the burden of different diseases in the communities studied, hence the importance of pooling efforts to overcome NTDs. The studies that have been conducted in Côte d’Ivoire (10, 31), Benin (61) and Burkina Faso(62), are examples of integrated approaches to NTD control, elimination and eradication. These studies have shown that integrated approaches can be effective in reducing the prevalence of NTDs and improving the health of people in these countries. Moreover, since many skin-NTDs are zoonotic or have a (possible) non-human reservoir, a One Health guided approach should be used to handle integrated case search activities (63). Synergies between skin-NTD programs and, for example, neglected zoonotic disease programs (such as rabies elimination) could optimize the use of resources (logistics) and stimulate cross-sectoral collaboration that is key for the sustainable control of NTDs.

This study has some limitations. The skin-NTDs reported apart from yaws were only suspected cases because the diagnoses were made based on clinical examinations. Unfortunately, the additional diagnostic tools required for confirmation of the clinical diagnosis were not available for this investigation. However, the data collected are still useful for guiding where skin-NTD eradication, elimination or control work should be directed in the future, i.e., data collection, training, diagnostics and the needs for medical care in these remote areas. We also acknowledge that the field visits were focused on areas where suspected yaws cases were reported which constitutes a bias for the estimation of other skin-NTDs.

This study highlights how the use of existing disease-specific resources can be maximized to determine the burden of other NTDs. Integration, technical and scientific progress are all pillars of the road map to end the suffering caused by NTDs by 2030.

## CONCLUSION

We demonstrate improved data reporting of skin-NTDs when an integrated approach is used. This study contributes to the strengthening of the yaws surveillance system into a guided integrated system that is being put in place throughout each country. The detection of yaws cases in areas that have undergone MDA is remarkable and highlights the importance of rigorous active surveillance ideally with test confirmation of clinically suspected cases and optimal coverage of MDA campaigns. The main challenges remain the difficulty of accessing some communities where yaws is endemic, the follow-up of patients, and the inter-community movements which are sources of reinfection, and resources to manage the affected populations.

## Data Availability

Data of this work are available through request to the corresponding author.

## ACKNOWLEDGEMENTS

This study is part of the Lamp4Yaws EDCTP funded project (grant number RIA2018D-2495). Our gratitude goes towards all participants and individuals involved in the field.

